# Development of a Core Outcome Set for Neurological Disorders (COS-Neuro): an AI-Assisted Thematic Framework Analysis

**DOI:** 10.1101/2025.09.22.25336392

**Authors:** Shyun Ping Tiong, Xiaoyu Yang, Alvaro Yanez Touzet, Christopher Paul Millward, Carl M. Zipser, Lindsay Tetreault, Ali Gharooni, Benjamin M. Davies

## Abstract

**Background:** Neurological disorders affect approximately 3 billion people globally, yet clinical trial success is often hindered by poorly selected outcome measures, impacting trial design, compliance, and interpretation. Over the past 25 years, Core Outcome Sets (COS) have emerged as standardized tools to enhance outcome selection, ensuring comparability across studies and reflecting the priorities of both researchers and patients. Despite the success of COS initiatives in other fields, their development in neurology remains limited, leaving many trialists without disease-specific guidance.

**Objectives:** This study aimed to develop a COS framework for neurological disorders with the assistance of artificial intelligence (AI) by analysing the frequency and scope of outcomes previously reported in existing COS to identify common themes applicable to neurological research.

**Methods:** COS-Neuro was developed using AI-assisted thematic framework analysis, complemented by expert review. A modified five-step thematic analysis was conducted without pre-determined codes:

1. Dataset Gathering – Data was collected from the COMET database, and COS domains for neurological disorders were coded.
2. Prompt Design & Testing – Large language models (LLMs), including ChatGPT 3.5, Google Gemini 1.5 Flash and Meta Llama-2-70b, were trialled, and prompts refined based on their outputs.
3. Thematic Analysis – LLMs categorised domains into core areas.
4. Human Refinement – Experts reviewed LLM-generated core areas and selected those most appropriate for further interpretation.
5. Clinical Validation – Experts validated the domains, core areas, and concepts.

This approach integrated AI with expert oversight to develop a standardised COS framework for neurological disorders.

**Results:** Utilising LLMs, particularly ChatGPT, a robust conceptual framework for COS in neurological disorders was developed, based on the existing 112 existing COS. Through adaptation of the OMERACT model, the final framework comprised four concepts, 13 core areas, and 75 domains, as determined by expert consensus.

**Conclusion:** COS-Neuro establishes AI-assisted framework for developing COS in neurological disorders. This project provides a foundational resource for future COS research and serves a reference for designing trials in areas where established COS are lacking. Furthermore, it sets a precedent for the integration of AI in qualitative analysis in medicine, demonstrating the scalability of approaches like OMERACT for the development of ‘COS of COS’ across various specialties.

## Introduction

Neurological disorders are the leading cause of illness and disability worldwide, affecting an estimated three billion people (1). Translational research is critical to addressing these unmet needs (2). As of March 2025, more than 10,800 clinical trials in neurological disorders were ongoing, accounting for 21.5% total clinical trials registered on clinicaltrials.gov.

While trial success depends on multiple factors, the choice of trial outcome measure is fundamental. Outcome measures inform sample size calculation, influence participant compliance and attrition, and ultimately determine how trial results are interpreted; put simply, if outcome measures are poorly chosen, a trial may reach incorrect conclusion or struggle to influence clinical practice (3–5).

An important initiative supporting outcome selection in clinical trials over the last 25 years has been the development of standardised outcome sets, known as Core Outcome Sets (COS) (6). COS are a list of critical disease features that should be measured for a particular disease or condition. Primarily designed to facilitate research synthesis by ensuring studies can be compared without bias, COS developed through broad stakeholder engagement - including individual lived experience - also help ensure that studies measure a disease comprehensively and capture outcomes that matter most to patients (7). Initiatives such as COMET (Core Outcome Measures in Effectiveness Trials) and OMERACT (Outcome Measures in Rheumatology) have significantly advanced COS development and refined best practice for their creation. A recent impact assessment of the COS for rheumatoid arthritis, one of the earliest examples, highlights the potential benefits for trialists, with translational success in the field correlating with its adoption rate, which now stands at 82% (8).

The potential for COS to support translational science is clear; however, their development and implementation remain time consuming and costly (9). To date, only 55 established COS (as of August 2024) have been developed for neurological diseases, including cases where multiple COS exist for a single condition within the COMET database. Consequently, many neurological trialists lack disease-specific COS as a reference when designing their trial (10). Although frameworks such as the OMERACT Filter provide conceptual guidance, they remain largely specific rheumatology and may not fully address the unique complexities of neurological research.

A review of neurological COS reveals many common themes, leading us to hypothesize that a shared framework - a ‘COS of COS’ - might be possible for neurological disorders. Unlike the OMERACT Filter, such sector-specific guidance could assist trialists working in conditions without established COS, as well as support COS developers in creating new disease-specific sets.

This study (COS-Neuro) describes the development of a COS framework for neurological disorders with the assistance of artificial intelligence (AI) by analysing the frequency and scope of outcomes previously reported in existing COS and generate common themes applicable across neurological research.

## Methods

This study employed a mixed-method design, combining semi-automated AI-based analysis with expert review to propose cross-cutting themes for neurological disorders.

Thematic analysis (TA) is a qualitative research method for identifying and analysing patterns or themes within a dataset through process such as data familiarisation and coding (11). AI is increasingly transforming qualitative research due to its cognitive ability in data processing, coding and pattern recognition. It has been shown to effectively assist in process of thematic analysis (12). Large Language Models (LLMs) are advanced AI models trained on massive text datasets, enabling them to comprehend contextual data. Coupled with natural language processing (NLP), LLMs can generate contextually relevant outputs and identify themes by analysing patterns across diverse domains (13). With the appropriate prompt, LLMs can enhance data visualisation, analysis and summarisation of scientific text (14, 15).

This COS-Neuro project was developed using thematic framework analysis (16). Themes were generated through AI-assisted identification and interpretation of data, followed by review and refinement by clinicians with experience and expertise in neurology and neurosurgery, COS and / or clinical trials.

A panel of six clinicians / researchers participated in the expert review and consensus process for COS-Neuro. Experts were purposively selected based on:

- Clinical experience in neurology or neurosurgery.
- Prior involvement in COS development or outcome research.
- Academic publications related to neurological disorders.

The panel included neurologists, neurosurgeons, and clinical researchers from the United Kingdom, Switzerland, and the United States. Table 1 summarises their professional background and relevant expertise.

**Table 1:**
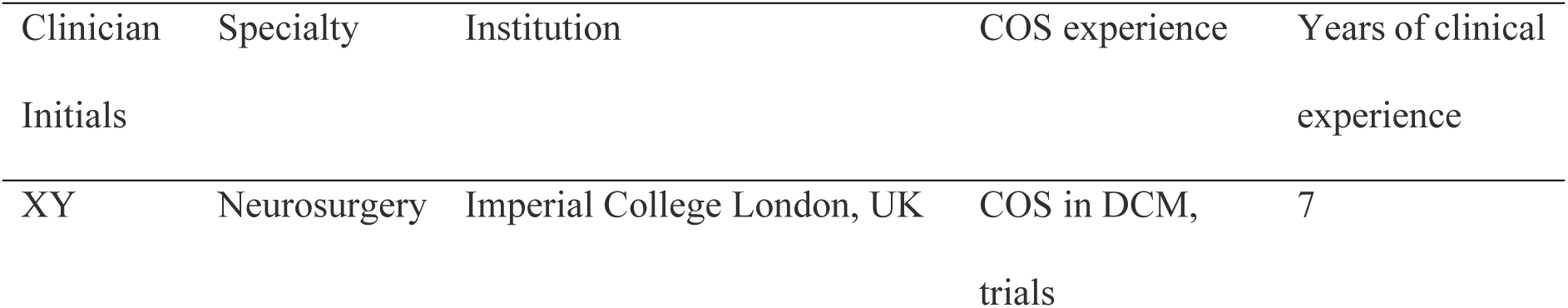
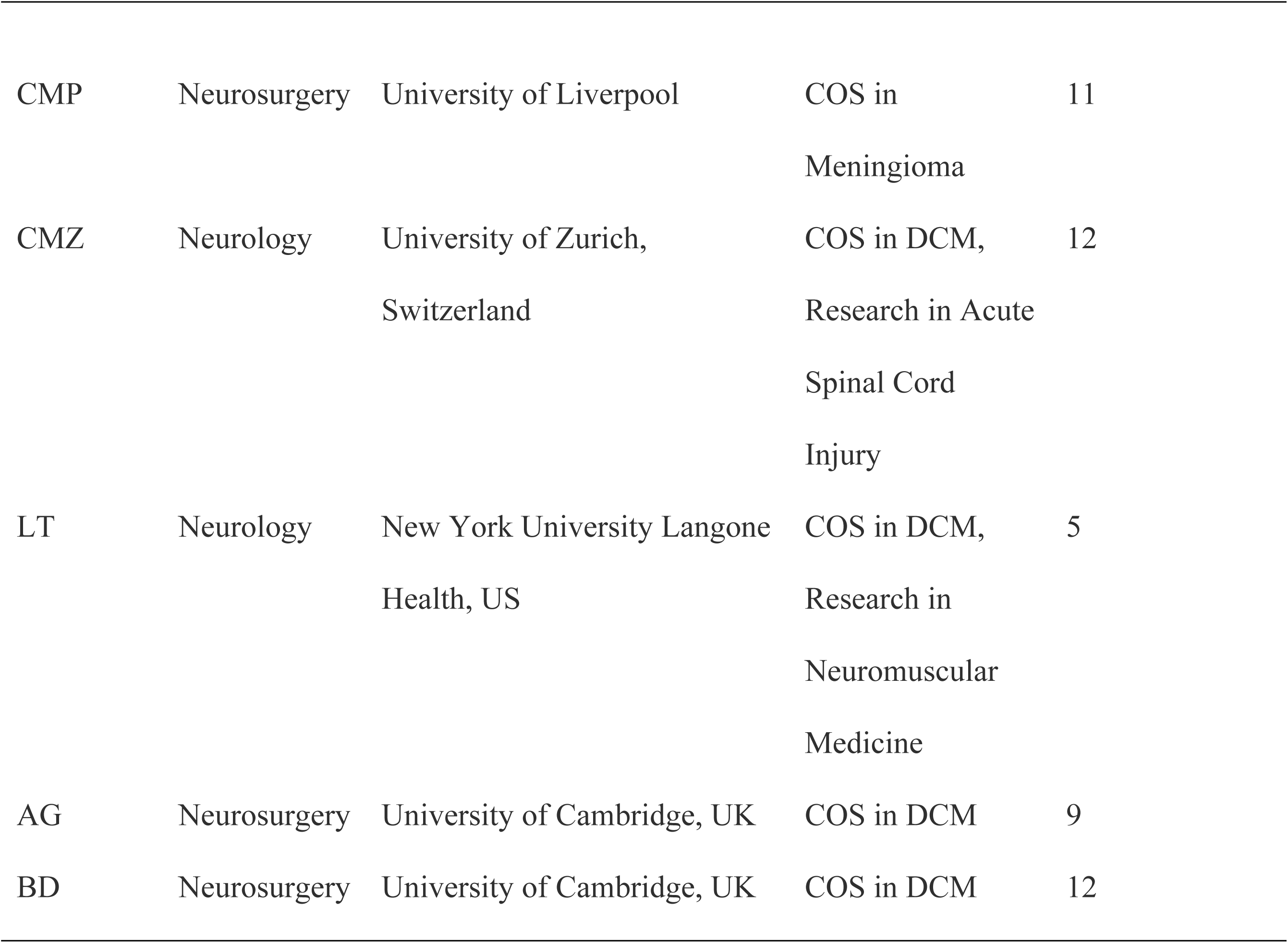
Background and Expertise of Panels Participating in Consensus Process.

| Clinician<br>Initials | Specialty | Institution | COS experience | Years of clinical<br>experience |
| --- | --- | --- | --- | --- |
| XY | Neurosurgery | Imperial College London, UK | COS in DCM,<br>trials | 7 |
| CMP | Neurosurgery | University of Liverpool | COS in<br>Meningioma | 11 |
| CMZ | Neurology | University of Zurich,<br>Switzerland | COS in DCM,<br>Research in Acute<br>Spinal Cord<br>Injury | 12 |
| LT | Neurology | New York University Langone<br>Health, US | COS in DCM,<br>Research in<br>Neuromuscular<br>Medicine | 5 |
| AG | Neurosurgery | University of Cambridge, UK | COS in DCM | 9 |
| BD | Neurosurgery | University of Cambridge, UK | COS in DCM | 12 |

The consensus process followed a modified nominal group technique. After initial AI-assisted thematic analysis, experts reviewed the proposed core areas and domains in an iterative process:

1. Round 1: Experts individually reviewed the initial AI-derived domains and provided written feedback via email.
2. Round 2: A virtual meeting was held where experts discussed discrepancies, suggested domain refinements, and proposed new domains or core areas.
3. Round 3: A revised list was circulated for final approval. Consensus was defined as ≥75% agreement on inclusion of domains or thematic groupings.

Disagreements were resolved through discussion during meetings until consensus was achieved. Detailed records of feedback and revisions were maintained to ensure transparency. Clinical validation thus integrated AI output with human expertise to establish a robust, clinically meaningful COS framework for neurological disorders.

Several initiatives have contributed significantly to the development and dissemination of COS, including OMERACT (17) and COMET initiative (18). Amongst the methodological resources created by OMERACT is the OMERACT Filter, which was updated in 2019 to Version 2.1. The filter aims to support COS developers in ensuring content validity by categorising the universe of measurable healthy-related concepts (19). The framework defines a measurement ‘concept’, such as Pathophysiology and Impact, which maps to a ‘Core Area’, such as Death, Life Impact, Societal/Resource Use, Manifestation and Abnormalities (Figure 1).

**Figure 1:**
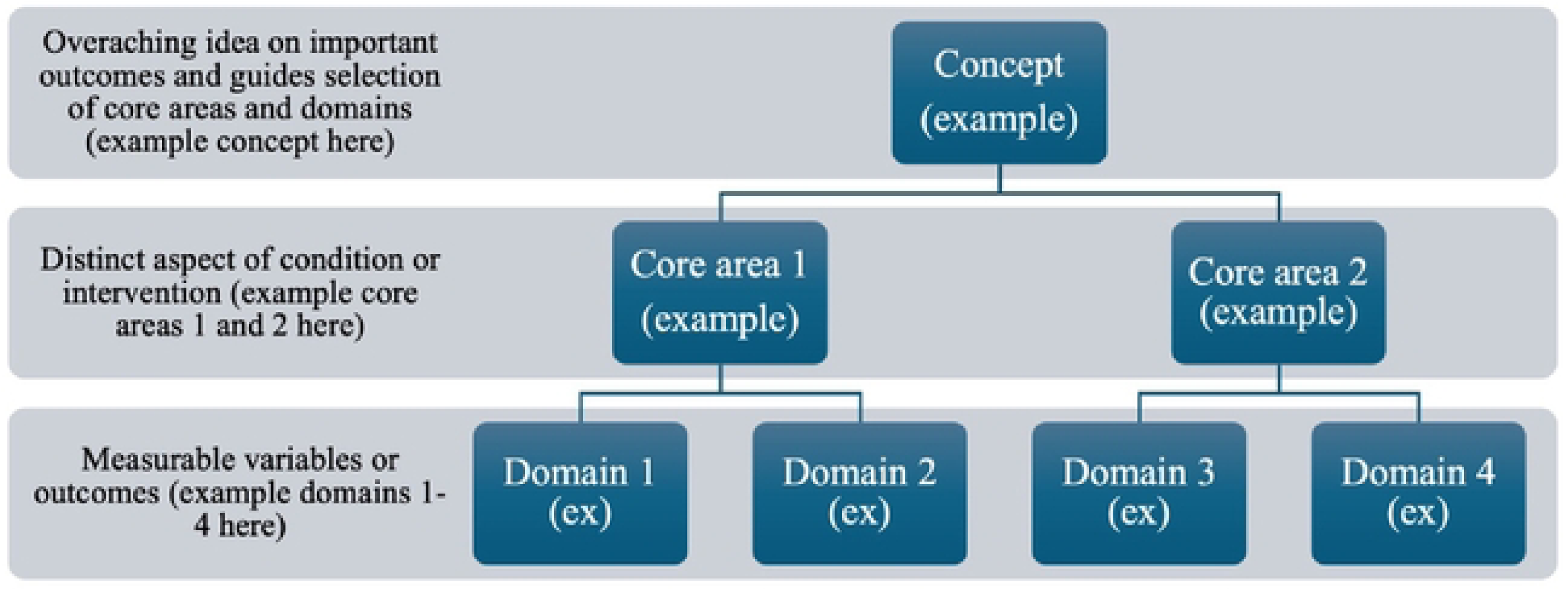
Hierarchical diagram for COS-Neuro development (adapted from OMERACT Filter 2.1)

Building on successes of OMERACT, the primary goal of COS-Neuro was to adapt OMERACT filter to develop a COS - of - COS framework for neurological disorders using AI-assisted thematic analysis combined with expert reviews. The development of COS-Neuro was pragmatic and intentionally provisional, prioritising efficiency amid uncertainty whether the approach was feasible. Therefore, the process relied on a small working group, with consideration of balance in expertise and geographic representation.

A modified five-step thematic analysis framework was used, where the themes were created without any predetermined codes (20).

### Step 1 - Dataset gathering

Data were extracted from the COMET database, which stored studies relevant to development of COS for use in clinical trials (21). The term ‘neurology’ was used in the search, and only published or ongoing studies were included in the dataset at the time of data collection (August 2022). The focus was on neurology, neurosurgery, and stroke as main specialties.

The COS were selected by reviewers with expertise in neurological disorders (neurosurgeons and neurologists) with inclusion and exclusion criteria outlined in Table 2. Each study was coded based on the COMET ID to facilitate cross-checking of domains by different reviewers.

**Table 2.** Inclusion and exclusion criteria for selecting existing Core Outcome Sets (COS)

| Inclusion criteria | Exclusion criteria |
| --- | --- |
| Neurology-related studies | Unpublished COS |
| Brain injury | Withdrawn COS |
| Spinal cord disorders | Ongoing COS |
| Dementia |  |
| Neurodegenerative diseases |  |
| Brain injury |  |
| Brain tumour |  |
| Functional neurological disorder |  |

Established core outcomes for each neurological disorder were longlisted in Microsoft Excel (Version 16.91) for analysis.

### Step 2 - Prompt design and trial

Three LLMs - CHATGPT 3.5, Google Gemini 1.5 Flash, and Meta Llama 2 70b - were used to analyse the longlist of domains. These models were selected due to their accessibility and advanced developed in the field of AI (22):

ChatGPT builds on Transformer Neural Network architecture, excelling in a wide range of NLP Task. It predicts the next word based on the contextual learning from pre-training and is capable of identifying statistical patterns and structural characteristics of human language. Google Gemini is known for outperforming human respondents in Massive Multitask Language Understanding (MMLU) tasks, including standardised exams and image recognition, through training on multimodal and multilingual datasets. Llama encompasses pre-trained and fine-tuned generative text models ranging from 7 billion to 70 billion parameter (22).

Prompt engineering was adapted to design prompts to guide thematic analysis using LLM. This involved optimising prompts to achieve desirable outcomes from generative model. In this study, instructions + input subgroup approach (23) was used to construct prompts across all three models.

Role prompting was employed as an effective strategy to initiate the task for a specialised topic (24). Prompts were structured on included a background information, specific instructions, and examples to minimise deviation from expected outcome.

For this project, prompts were framed to assign the LLM the role as medical researcher (a trialists), with instructions to perform thematic analysis on core outcome measurements and propose suitable cross cutting themes based on given domains.

A stop criterion for further prompt generation was defined as thematic saturation - when all the major domains of neurological disorder outcomes were adequately represented in the core set at clinician’s consensus, and additional analysis no longer yielded new insights (25).

Response across the three LLMs was reviewed, with feedback incorporated for further refinement.

### Step 3 - Thematic analysis

An in-depth inductive thematic analysis was conducted to generate outcomes domains. Data were analysed inductively using a combination of LLMs outputs and clinical expertise to identify and interpret outcomes relevant for patients (26), and to assess the quality of the dataset used in themes generation.

Domains considered broadly relevant across all neurological disorders were identified. Under clinician oversight, outputs from each model were compared and annotated for further improvement.

### Step 4 - Refining output with human expertise, and selection of preferred LLM

Each LLMs were prompted to provide rationales for its outputs. Output was then compared, and the most suitable LLM was selected by clinicians for further analysis.

The selection criteria included:

1. The ability to accept long prompts sufficient to process the entire domain longlist
2. Generation of outputs that accurately linked domains to relevant core areas
3. Core areas deemed acceptable and meaningful by clinicians

Once the core areas were finalised, a new prompt was designed and tested to link domains to core areas in a tabular form, which was subsequently extracted into Microsoft Excel.

### Step 5- Hierarchical categorisation using clinical expertise, and OMERACT Filter 2.1

Clinicians worked collaboratively, using the OMERACT Filter 2.1 as a reference framework, to aggregate core areas, into high-level categories (Figure 1) (27). This included refining the terminology used to describe each core area generated by LLM. Experts were encouraged to draw on their broader medical experience, rather than simply rely on the outcomes from existing neurological COS, to ensure that the resulting framework could be generalised across neurological disorders.

## Results

### Step 1 - Dataset gathering

A total of 112 articles on COS relevant to neurological disorders were identified on COMET database, from which 782 core outcomes were extracted. Each study was coded with the COMET ID, and the outcomes were compiled into longlist based on these ID (Supplement 1).

There were 55 neurological disorders with established COS, including condition for which multiple COS exist.

### Step 2 & 3 - Prompt design and initial thematic analysis

The initial prompt was designed to instruct LLMs to perform thematic analysis based on the given longlist of domains. LLMs were specifically asked to provide themes with examples for analysis of the thought process.

Prompt 1:

> *Act as a medical researcher. You are given a longlist of core outcome measurement sets on neurology. Please do an inductive thematic analysis of the longlist and summarise maximum of 20 themes. I want domains as examples that linked to the themes. Example of themes would be comprehensive health assessment with symptoms, blood pressure, mental health as domains.*

> *\*Longlist of outcomes\**

Feedback on outcome for prompt 1

ChatGPT and Gemini both focused on symptoms, health status, and quality of life, which were subsequently selected as the main themes. The prompt length was too extensive for Llama, necessitating further refinement. The prompt was improved by incorporating common themes reported by both ChatGPT and Gemini as examples and was slightly shortened to accommodate Llama’s input limitations.

The prompt was further edited and improved based on feedback until satisfactory output was achieved in prompt 6. Details on the prompts and their outputs are provided in Supplement 2.

Prompt 6:

> *Act as a medical researcher. You are given a longlist of core outcome measurement sets in different neurology disorders. please do an inductive thematic analysis of the given longlist and summarise the main themes suitable for COS in neurology. Please use OMERACT as example with concepts, core areas and domains.*

> *\*Longlist of outcomes\**

The outcome generated by ChatGPT is provided in Supplement 3.

### Step 4 - Refining output with human expertise, and selection of preferred LLM

Outputs from ChatGPT, Gemini and Meta were assessed by clinicians based on the criteria detailed in the Methods section. ChatGPT effectively summarised key points relevant in neurology. Gemini followed the OMERACT framework and produced concepts similar to ChatGPT, although its examples were less relevant to neurological disorders. Llama was considered unsuitable for this purpose, as it was unable to process a long list of domains or provide coherent justification for its output. Clinicians therefore agreed to proceed with the outputs of ChatGPT from prompt 6 (Supplement 4) as the foundation for further COS development.

ChatGPT generated 10 themes, which were then established as foundation for COS-Neuro development: Disease Activity/Progression, Physical Functions, Quality of Life and Patient-reported Outcomes, Neurological Symptoms, Health Economics and Health Services, Functional Neurological Disorder (FND) Symptoms, Rehabilitation Outcomes, Perioperative and Surgical Outcomes, Paediatric Neurology Outcomes and Visual and Ocular Outcomes.

ChatGPT was subsequently prompted to classify and tabulate all the relevant domains into these themes. Two rounds of prompt were used to successfully reduce 782 total domains to 225 domains considered relevant to all neurological disorders. ChatGPT organised the domains into themes/core areas in a table (Supplement 5). With clinicians’ oversight, the domains were re-categorised or removed based on clinical relevance.

Prompt 2:

> *“Please analyse ALL the domains given and classify them into the above themes. Provide your answer in a table with two columns. I want the first column to be ALL the longlist of domains given in the first prompt, second column to be the themes. The themes are disease activity/progression, physical functions, Quality of Life and Patient-reported Outcomes, Neurological Symptoms, Health Economics and Health Services, Functional Neurological Disorder (FND) Symptoms, Rehabilitation Outcomes, Perioperative and Surgical Outcomes, Paediatric Neurology Outcomes and Visual and Ocular Outcomes”*

### Step 5 - Hierarchical Categorisation using Clinical Expertise, and OMERACT Filter 2.1

Table 3 outlines the rationale behind each modification to the core areas, including rewording, combining, and further sub-categorisation based on consensus process.

**Table 3:** Improvement of core areas during consensus.

| **Initial core area** | **Improved core area** | **Rationale** |
| --- | --- | --- |
| Disease activity/progression | Disease-specific activity/progression outcome | The domains in this category focused primarily on seizure activity and control, so disease-specific activity was more appropriate. |
| Neurological symptoms<br>Visual and ocular outcomes | Pain<br>Sensory function<br>Vision<br>Autonomic functions<br>Cognition<br>Motor functions | Neurological symptoms and visual/ocular outcomes were too broad as core areas, so it was sub-categorising into different neurological functions |
| Health economics and health service | Burden of disease and economic impact | It was rephrased to healthcare delivery as the domains also consisted of adherence, medication appropriateness, compliance to treatment, education/support which were not limited to economics and services |
| Quality of life | Quality of life<br>ADL<br>Coping<br>Social participation | QoL had been sub-categorising into QoL, ADL, Coping and social participation as the domains covered a wide range of patient-reported outcomes which were vital in neurological disorders |
| Rehabilitation outcomes<br>Perioperative and surgical outcomes | Treatment process and outcomes | The two core areas were combined into treatment process and outcomes as they were interconnected and collectively influenced the treatment recovery. |
| Paediatric neurology outcome | Child development and formation | Paediatric might be easily misunderstood as mixing up with another speciality and the domains such as preschool attendance and performance, engagement in school life were more holistic approach into children's development |
| Functional neurological symptom disorder | Removed | FND is a diagnosis rather than a core area. |

The final core areas agreed upon by clinicians were disease specific activity, Pain, Sensory function, Vision, Autonomic functions, Cognition, Motor functions, Burden of disease and economic impact, Quality of life, ADL, Coping, Social participation, Treatment process, and Child development and formation.

Once these core areas were established, a final round of prompts was designed to aggregate LLM-generated domains into overarching themes, referred to as ‘main domains’. The aim was to create descriptive terms that would intuitively prompt COS-Neuro users to understand the construct of each area.

Final round of prompt to identify main domains:

> *‘Act as medical researcher. You’re developing core outcome measurements in neurological disorders. You’re given the main core areas followed by their domains as text below. I want you to identify the MAIN domains for the core areas.’*Through this process. 225 domains were further consolidated into 75 domains, as tabulated in Table 4.

**Table 4:**
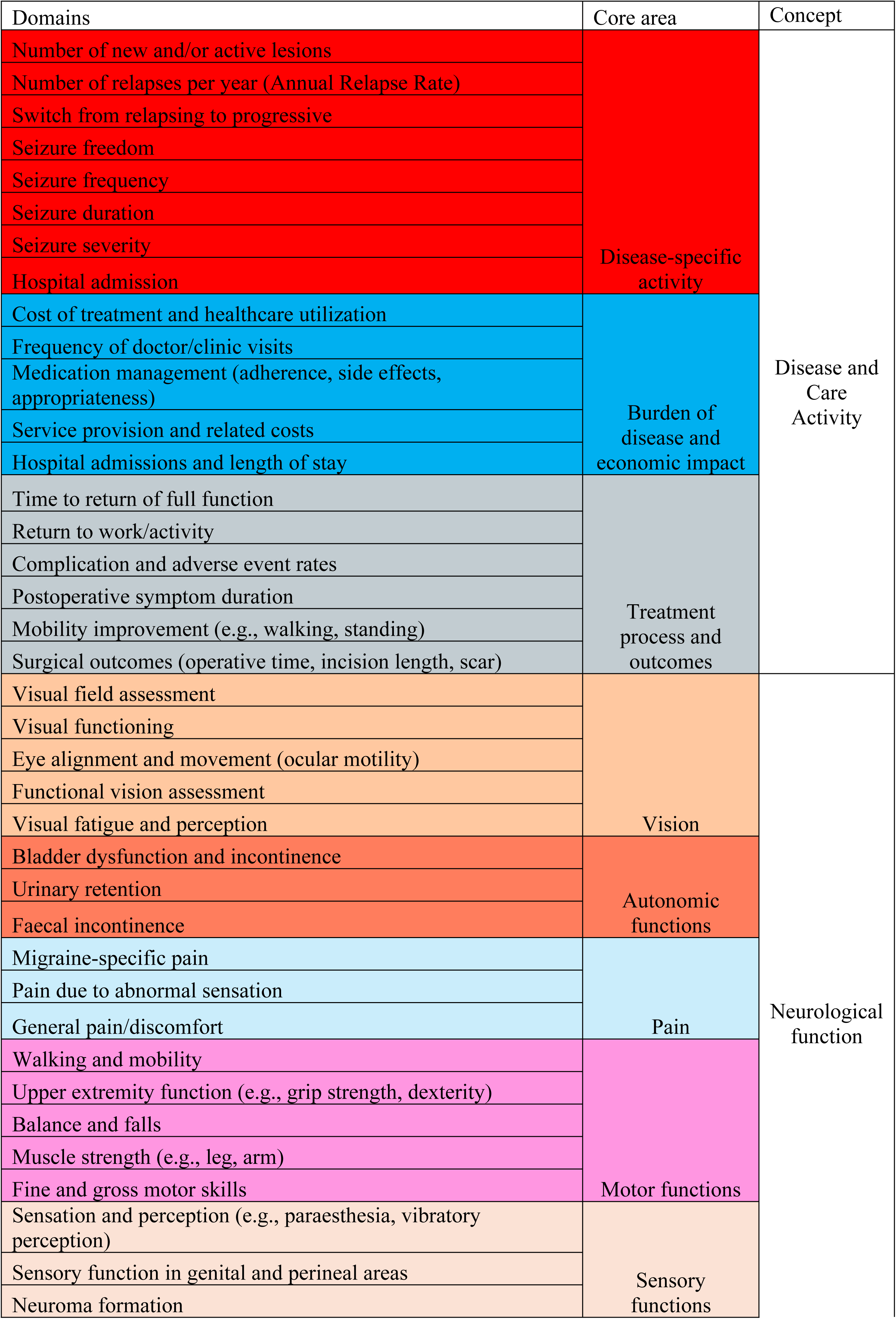
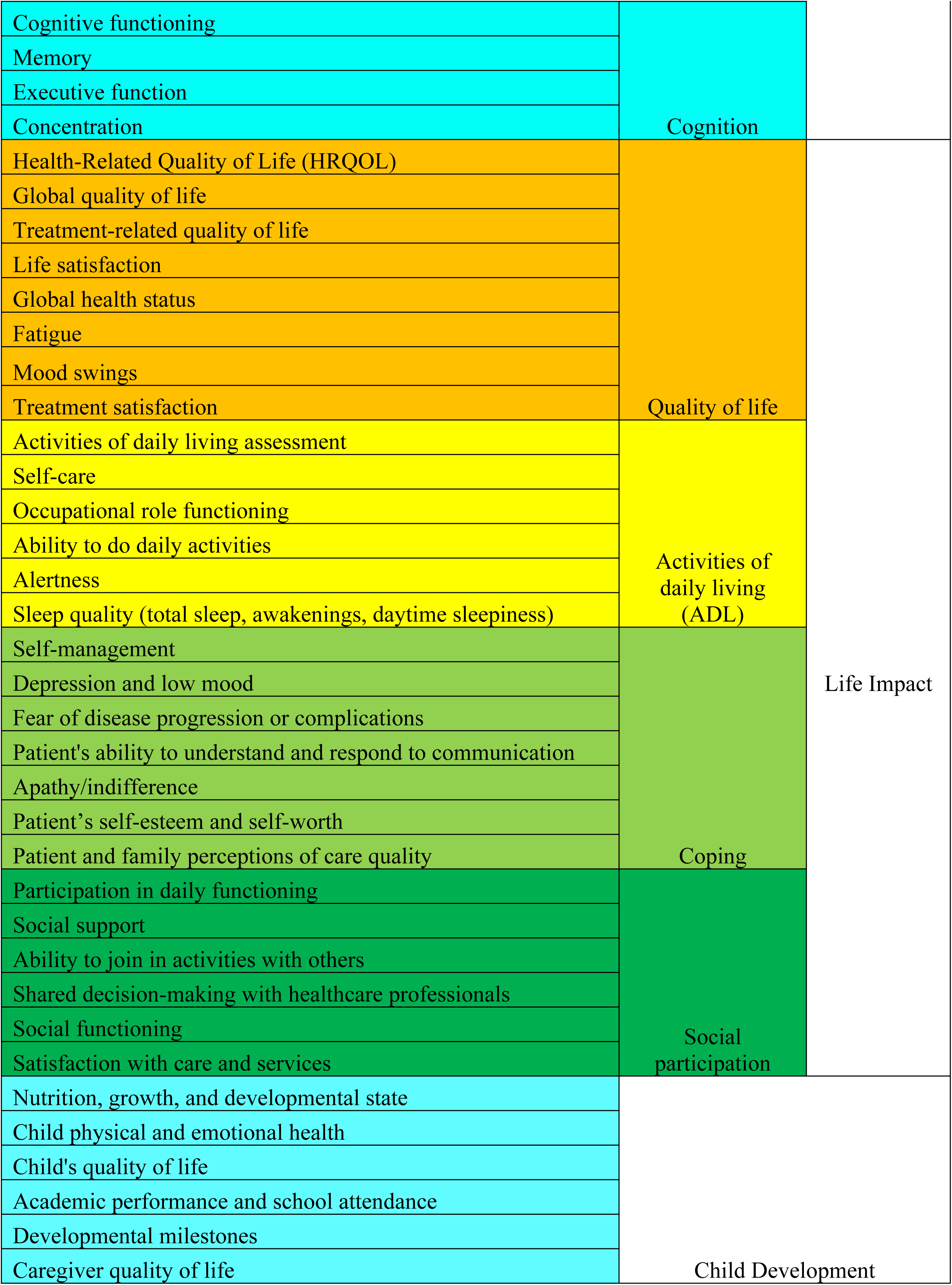
AI-identified main domains based on clinicians agreed core areas and concept.

The final framework for COS-Neuro included four main concepts as agreed in consensus: Life impact: including quality of life, ADL, coping, and social participation, representing the social aspects of patients’ life living with neurological disorders.

Neurological function: including vision, autonomic function, pain, motor, sensory and cognition, to establish baseline for patients and characterise disease features.

Disease and care manifestations: sub-divided into disease-specific activity, burden of disease and economic impact, treatment process and outcomes, aimed at evaluating disease progression and therapeutic effectiveness.

Child development and formation: an optional fifth concept, included to address specialised outcomes in paediatric neurological disorders where relevant.

The final framework for COS-Neuro is illustrated in Figure 2.

**Figure 2.**
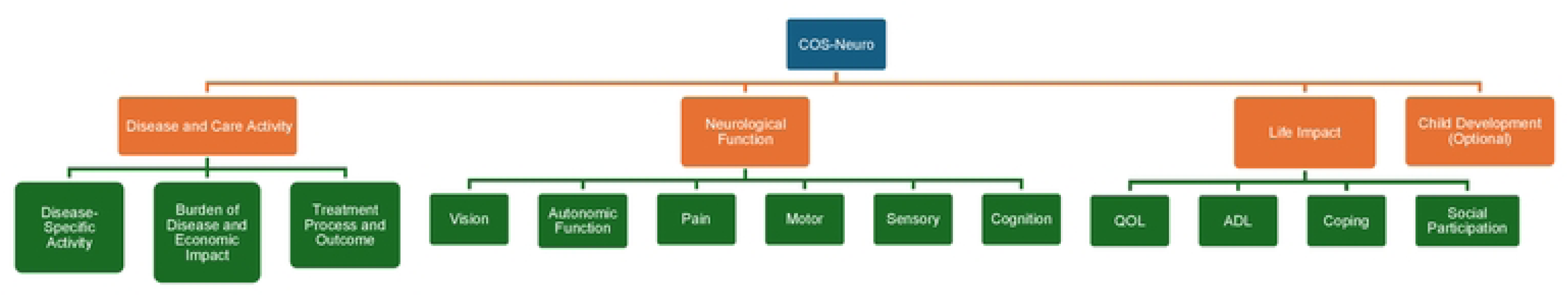
Final framework for COS-Neuro. Four concepts (Orange) were identified; *Disease and Care Activity, Neurological Function, Life Impact and Child Development.* These mapped to 13 Core Areas (Green). *QOL* Quality of Life. *ADL* Activities of Daily Living.

## Discussion

There is growing awareness and emphasis on using COS in the design and conduct of neurological clinical trials. This is evidenced by their increasing adoption (47% for clinical trials (28)), and ongoing development (48 COS currently in planning or ongoing, according to the COMET database on 26/03/2024)). Nevertheless, many neurological conditions remain unrepresented.

With the assistance of LLMs, particularly ChatGPT, a robust conceptual framework for COS in neurological disorders was developed, based on the existing 112 existing COS in the COMET database. A total of four concepts, 13 core areas and 75 domains were finalised following clinical consensus. Given the heterogeneous presentation of neurological diseases, 75 domains were identified to ensure comprehensive coverage of relevant aspects of disease impact. The concept of functional impairment facilitates the evaluation of clinical progression and therapeutic effectiveness. Neurological function domains help establish baselines for patients and understand disease characteristics, while patient-centred outcomes capture the social impact of neurological disorders. Child development and formation were treated as a distinct, optimal concept/core for paediatric neurological disorders.

To our knowledge, COS-Neuro is the first project to develop a ‘COS of COS’ for neurological disorders, representing a significant methodological advancement in the COS and clinical research by integrating AI into thematic analysis. A recent study (29) has demonstrated ChatGPT’s ability to enhance the efficiency in thematic analysis by generating meaningful sets of codes and themes.

Traditional COS development typically requires manual qualitative data analysis, which is time-consuming and labour-intensive, due to large volumes of textual data that needs to be explored inductively to generate themes and categories (30). The development of Computer Assisted Qualitative Software (CAQDAS) - such as MAXQDA, NVivo, ATLAS.Ti, and N6-marked an important milestone in qualitative research. These tools offer features such as character-based coding, rich text capabilities, and multimedia support. While CAQDAS improves data transcription efficiency and accelerate analysis, it is underutilised among qualitative health researchers (reported use of only 16%), partially due to the complexity of mastering the platform (31). Additionally, they are costly, with MAXQDA priced at up to €220 yearly for single user (as of March 2025).

By contrast, LLMs offer the ability to rapidly process large datasets, identify patterns, generate themes, and summarise key findings - significantly reducing the time and resources required for COS development (32). Most LLMs are free to access and communicate with plain text, making them more intuitive than traditional software. However, effective prompt engineering is pivotal for ensuring relevant, informative, and accurate AI-generated responses. Well-crafted prompts significantly enhance the performance of AI models (23). In this study, all prompts were carefully designed and under clinical oversight to optimise the outputs. Collaborations between human and LLMs are crucial in improving consistency and reliability of AI-integrated thematic analysis (33).

### Advantages and Challenges of AI integration

The integration of AI into thematic analysis offered several advantages:

*Efficiency*: AI significantly reduced the time required to analyse 782 domains extracted from 112 COS studies, generating 75 meaningful domains relevant to the core areas.

*Consistency*: ChatGPT demonstrated high consistency in identifying common themes and providing rationale, improving inter-rater reliability.

*Scalability*: This study establishes a replicable and scalable approach for AI-assisted COS development, which can be extended beyond neurology to other specialities.

Although AI has been used in medical research (29, 34), this is the first know application of AI-assisted thematic framework for COS development. Previous studies have demonstrated the utility of ChatGPT as an assistive tool for improving the efficiency of qualitative research, but none have used AI to develop COS from a longlist of domains.

Nevertheless. AI integration poses several challenges. A major limitation is the variability in output between different LLMs, even when using the same dataset and prompt. This is due to differences in their underlying architectures and training datasets which may contain bias(32, 35). To mitigate this, we used the same prompt across three models. ChatGPT was selected for final analysis based on its superior consistency and domain relevance, as determined by clinical expertise.

Another key issue is the phenomenon of AI ‘hallucinations’ – where the model generates inaccurate, misleading, or fabricated content. This was addressed by using structured prompt engineering and continuous refinement (36). A clinician was assigned to lead the process, assess model outputs, and verify the validity of the results. LLMs were also prompted to explain their reasoning, enabling human reviewers to evaluate and, where necessary, correct thematic interpretations. This human-AI collaboration ensured that the final COS-Neuro framework reflect real-world clinical priorities (37).

### Limitations, Future Development and Implementation

While COS-Neuro represents a significant step towards standardisation of outcome selection in neurological trials, several limitations were identified. The datasets were based exclusively on the 112 existing COS for neurological disorders in the COMET database. As such, the framework reflects the limitation of those existing COS and may not fully capture the complexity of all neurological conditions. Several important aspects of neurology, including sleep and behavioural symptoms such as hallucinations and delusions, should be considered in developing COS, as recognised by consensus among clinical experts given their common presentation in neurological diseases. However, these domains or core areas have not been established as a COS and were therefore not included in the considerations in COS development.

Development of COS often involves a broad consensus group with various stakeholders to ensure meaningful and relevant outcomes. However, the development of COS-Neuro was deliberately a streamlined process but considered as appropriate for several reasons: (a) Development of the existing COS in neurological disorders included in this study were inherently resource-intensive, (b) This approach is preliminary and subject to prospective validation and refinement, and (c) Our aim at this stage is to explore feasibility/efficiency.

The decision to adapt the OMERACT Filter was pragmatic. Like rheumatological conditions, neurological disorders are complex and heterogeneous. There is considerable overlap, e.g. neurological manifestations of vasculitis or connective tissue diseases. The OMERACT Filter was originally developed to improve content validity in outcome selection for Rheumatological trials and has been refined and adopted over time. It is also recommended in COMET Handbook as a recourse for COS development (7, 8, 38).

Looking ahead, ongoing improvement in AI models could further enhance COS-Neuro. Future iterations may incorporate underrepresented neurological areas, expand domain definitions, and refine thematic structure with additional stakeholder input.

### Practical implication

COS-Neuro can already serve as a conceptual framework to support the development of new COS for other neurological disorders. In the authors’ experience developing a COS for Degenerative Cervical Myelopathy, consolidating and categorising a large and diverse set of outcomes posed substantial challenges (39). A framework like COS-Neuro would have greatly supported the process.

COS-Neuro may also assist trialists in selecting outcome measures for new studies, particularly where no standardised endpoint exists. By providing a structured framework, it encourages the use of clinically relevant and patient-centred outcomes, thereby improving consistency and comparability across studies (7, 40).

## Conclusion

A conceptual framework to inform the selection of outcome measures in neurological trials and the development of neurological COS has been created with assistance from LLM, based on existing neurological COS and the OMERACT Filter 2.1. Of the LLM explored, ChatGPT was considered to be the most effective for this task. The semi-automated nature of this process can support future iterations of COS-Neuro, and its application in other diseases, providing a blueprint for the development of ‘COS of COS’ framework.

## Data Availability

All relevant data are within the manuscript and its Supporting Information files.

## Acknowledgments

We are grateful to Mr Jeffrey Coates (Ex-Military Officer) for his assistance with prompt engineering.

We used generative AI tools — ChatGPT (41), Gemini 1.5 Flash (42), and LLaMA 2 70B (43) — to assist in thematic framework analysis.

## Authors Contribution

Conceptualization: SPT, XY, AYT CPM, CMZ, LT, AG, BMD

Data Curation: CPM

Methodology: SPT, XY, AYT CPM, CMZ, LT, AG, BMD

Writing-Original Draft and Editing: SPT, XY, AYT CPM, CMZ, LT, AG, BMD

## Abbreviations

| Acronym | Definition |  |
| --- | --- | --- |
| COS-Neuro | Core Outcome Sets in Neurological disorders | An ‘COS of COS’ initiative supporting the development of consensus in outcome measurement for neurological disorders. |
| COS | Core Outcome Sets | An agreed standardised set of outcomes that should be measured and reported, as a minimum, in all clinical trials in specific areas of health or health care. |
| OMERACT | Outcome Measures in Rheumatoid Arthritis Clinical Trials | www.omeract.org—An initiative supporting the development of consensus in outcome measurement for arthritis. |
| COMET | Core Outcome Measures in Effectiveness Trials | https://www.comet-initiative.org - An Initiative in development and application of agreed standardised sets of outcomes (COS) |
| TA | Thematic Analysis | Research method used to identify and interpret patterns or themes in a data set |
| AI | Artificial Intelligence | Technology that enables computers and machines to simulate human learning, comprehension, problem solving, decision making, creativity and autonomy |
| LLM | Large Language Models | A type of artificial intelligence algorithm that uses deep learning techniques and massively large data sets to understand, summarize, generate and predict new content |
| Concept | Overarching idea on important outcomes and guides selection of core areas and domains |  |
| Core area | Distinct aspect of condition or intervention |  |
| Domain | Measurable variables or outcomes from COS |  |
| Outcome | A measurable change or result that can be used to assess the effect of a health care intervention, condition, or disease |  |
| OMERACT Framework | A conceptual framework that helps identify the key areas and domains of health that need to be measured, ensuring a comprehensive approach to outcome measurement |  |
| OMERACT Filter | A tool used to assess and select the appropriate outcome measurement instruments (questionnaires, scales, etc.) for inclusion in the core outcome set. |  |

